# Patient-Tailored Minimally Invasive Hybrid Ablation of Complex Ventricular Tachycardia Substrates

**DOI:** 10.1101/2025.11.10.25339388

**Authors:** Rachel M.A. ter Bekke, Yesim S. Kaya, Laurent Pison, Reinder Evertz, Kevin Vernooy, Justin G.L.M. Luermans, Mindy Vroomen, Jens Jeurissen, Sevasti-Maria Chaldoupi, Elham Bidar, Roberto Lorusso, Barthel Sauren, Mark La Meir, Bart Maesen

## Abstract

**Background:** Clinical experience with minimally invasive hybrid ventricular tachycardia (VT) ablation remains limited, particularly regarding video-assisted thoracoscopic (VATS) access.

**Objectives:** To describe procedural characteristics, feasibility, and outcomes of minimally invasive hybrid VT ablation for complex substrates.

**Methods:** Consecutive patients undergoing minimally invasive hybrid VT ablation at a single tertiary center (2014–2025) were retrospectively analyzed. Multidisciplinary preprocedural evaluation was consistently performed, and from 2022 onward, this was formalized within the VT-TRACT (ventricular tachyarrhythmias: a multidisciplinary clinical-translational approach) care pathway.

**Results:** Twenty-two patients (86% male, median age 70 years, median PAINESD score 13, 68% VT storm) underwent minimally invasive hybrid VT ablation: left- or right-sided VATS (n=13 vs 1), subxiphoid access (n=3), double access (n=2), and anterolateral minithoracotomy (n=3). Indications were prior cardiac surgery in 7 (32%), extensive scar in 3 (14%), concomitant left-sided sympathectomy in 2 (9%) and hybrid atrial fibrillation ablation in 1 (5%), failed epicardial access in 2 (9%), pericarditis/tamponade in 2 (9%), while 5 (23%) underwent ablation under direct visualization by preference. Pericardial adhesions (45%) were bluntly dissected. Mean procedure time was 312±98 minutes. At one year, median VT burden decreased from 16.5 [9.5–37.0] to 0 [0–5.8] (−81%, *P*<0.001), and ICD shocks from 2 [0–5] to 0 [0–0] (−90%, *P*<0.001). One hemothorax required reoperation; no other major complications occurred. One-year survival was 82%.

**Conclusions:** Minimally invasive, patient-tailored hybrid VT ablation—guided by multidisciplinary planning—achieves marked reductions in VT burden and ICD shocks with a favorable safety profile, even in complex post-surgical patients.

## Introduction

Scar-related ventricular tachyarrhythmias (VT) remain a major cause of morbidity and mortality across cardiac etiologies. Although implantable cardioverter-defibrillators (ICDs) constitute the cornerstone of secondary prevention, they do not prevent arrhythmia recurrences, and repetitive shocks significantly impair quality of life while increasing mortality.^1,2^ Catheter-based VT ablation effectively reduces arrhythmia burden and ICD therapies, demonstrating superior efficacy compared with antiarrhythmic drug therapy.^3^

The arrhythmogenic substrate often exhibits complex three-dimensional architecture, involving the epicardium, particularly in nonischemic and arrhythmogenic cardiomyopathies, as well as in a substantial proportion of ischemic cases.^4^ Consequently, epicardial access and ablation are frequently required to achieve complete substrate elimination. Combined endocardial-epicardial ablation has been shown to improve long-term outcomes, reducing VT recurrence by approximately 35% compared with endocardial-only approaches.^5^ However, percutaneous epicardial access carries procedural risks (1–15%)^6^ and can be restricted by adhesions, obesity, or proximity to critical structures such as coronary arteries and the phrenic nerve.^7^ The presence of dense pericardial adhesions—common after cardiac surgery, prior epicardial ablation, or pericardial inflammation—may further preclude safe and effective percutaneous access.

To overcome these limitations, hybrid surgical–catheter approaches have been developed, combining limited surgical epicardial access via subxiphoid or anterior thoracotomy with mapping-guided ablation, allowing controlled adhesiolysis and direct visualization of the arrhythmogenic substrate.^8^ Although effective, these procedures are associated with long procedural times and significant complication rates in up to 20% of cases.^9^ Minimally invasive video-assisted thoracoscopic (VATS) hybrid ablation represents a promising alternative, enabling targeted substrate modification with reduced surgical trauma.^10^

Here, we present our 10-year single-center experience with minimally invasive hybrid VT ablation, predominantly using a left-sided VATS approach, demonstrating procedural feasibility, safety, and efficacy in a high-risk cohort with advanced structural heart disease.

## Methods

### Study Population

Consecutive patients with structural heart disease who underwent minimally invasive hybrid mapping and ablation of VT at Maastricht University Medical Center+ between 2014 and 2025 were included. Data were retrospectively analyzed as part of the Maastricht Multicenter Cardiology Monitoring Platform Study or “ventricular tachyarrhythmias: a multidisciplinary clinical-translational approach“ (VT-TRACT) registry (approved by the MUMC+/UM Ethics Committee; NCT04976348). Baseline demographic, procedural, and follow-up data were retrieved from electronic medical records.

### Preprocedural Evaluation and VT-TRACT Pathway

Multidisciplinary preprocedural evaluation was routinely conducted for all cases with recurrent VT and suspected epicardial substrate—based on ECG characteristics, prior endocardial ablation, or imaging findings. Beginning in 2022, this process was standardized and integrated into the structured VT-TRACT care pathway. The VT-TRACT team includes electrophysiologists, cardiothoracic surgeons with electrophysiologic expertise, interventional cardiologists, genetic cardiologists, and cardiac anesthesiologists. Each case was reviewed for relevant medical history, prior cardiac surgery, arrhythmia mechanism, electroanatomic maps (EAM) from previous ablations, and three-dimensional cardiac anatomy derived from cardiac computed tomography angiography (CCTA)^11^ and/or magnetic resonance imaging (CMR)^12^ to assess potential corridors, the trajectory of epicardial vessels, and the phrenic nerve before determining the interventional strategy. The PAINESD score was calculated to estimate the risk of periprocedural hemodynamic decompensation and guide decisions for preemptive veno-arterial extracorporeal membrane oxygenation support (V-A ECMO).^13^

Procedural indications included:^9^

1. History of prior epicardial or cardiac surgical interventions,
2. Failed percutaneous epicardial access for VT ablation,
3. Substantial transmural or epicardial substrate requiring extensive ablation,
4. Concomitant sympathetic denervation or hybrid atrial fibrillation ablation,
5. Prior tamponade, pericarditis,
6. Preferred ablation under direct vision.

Patients with severe pulmonary disease were excluded.

### Minimally Invasive Epicardial Access

Minimally invasive epicardial access was primarily achieved through a left- or right-sided VATS approach, depending on substrate location. In selected cases, a subxiphoid or anterolateral mini-thoracotomy was used (Figure 1, Panels A and B). All procedures were conducted in a dedicated hybrid electrophysiology (EP) operating room equipped with fluoroscopy, EAM systems, high-resolution surgical imaging, and facilities for immediate cardiopulmonary bypass or V-A ECMO support.

**Figure 1.**
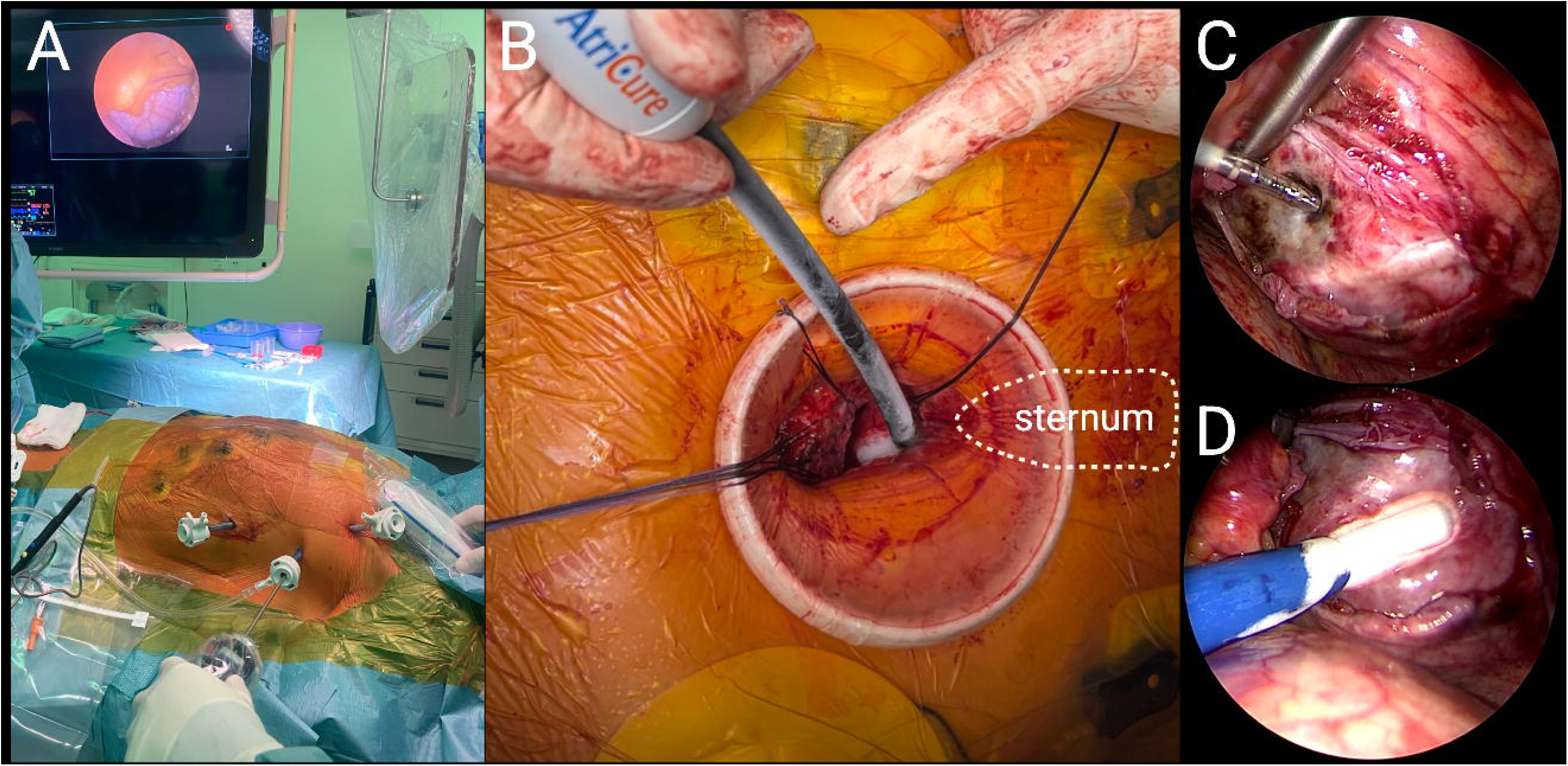
Minimally invasive hybrid VT access and ablation. In panel A, a left-sided video-assisted thoracoscopic approach through three 5-mm ports into the 3^rd^, 5^th^, and 7^th^ intercostal spaces permits access for surgical tools, electrophysiological mapping, and ablation catheters. In panel B, a surgical subxiphoid window with a soft-tissue retractor allows maximal epicardial exposure for cryo-ablation. Thoracoscopic radiofrequency (Panel C) and cryo-ablation *a vue* (Panel D), distal and proximal to an arterial bypass graft.

Patients were positioned supine with an inflatable cushion under the left or right shoulder. After positioning of the CARTO and defibrillation pads and initiation of continuous 12-lead ECG monitoring, general anesthesia was induced, followed by placement of a double-lumen endotracheal tube. The surgical field was prepared to allow simultaneous groin, subxiphoid, and thoracic access. All patients received prophylactic antibiotics.

### Video-Assisted Thoracoscopy

In most patients, a left-sided thoracoscopic approach provided optimal access to the left and right ventricular epicardium (Figure 1, Panel A, Central Illustration). After establishing single-lung ventilation, three 5-mm ports were inserted into the 3rd, 5th, and 7th intercostal spaces, triangulated around the anterior axillary line, guided by preoperative imaging.

A small pericardial incision was made, and the mapping catheter was introduced via a 5-mm trocar for initial mapping with the pericardium intact. Once mapping was complete, inital ablation was performed, after which the pericardium was opened around the target region for detailed mapping and ablation. Energy delivery was performed using a standard endocardial radiofrequency (RF) catheter, a linear bipolar RF catheter (Coolrail, Atricure, OH, USA), or a flexible cryoablation probe (Cryo, Atricure, OH, USA) (Figure 1, Panels C and D).

Thoracoscopy enabled direct visualization of coronary arteries and ventricular anatomy, controlled adhesiolysis, phrenic nerve displacement, and stable catheter positioning. It also enabled concomitant left cardiac sympathetic denervation (LCSD, Figure 2). For LCSD, the parietal pleura was opened over the stellate ganglion, from the distal half of C8 to T5 (Figure 2, Supplemental Video 1). The sympathetic chain was divided below the stellate ganglion and clipped, sparing the branch to the left arm. All side branches were transected, and the excised segment was submitted for histologic confirmation.

**Figure 2.**
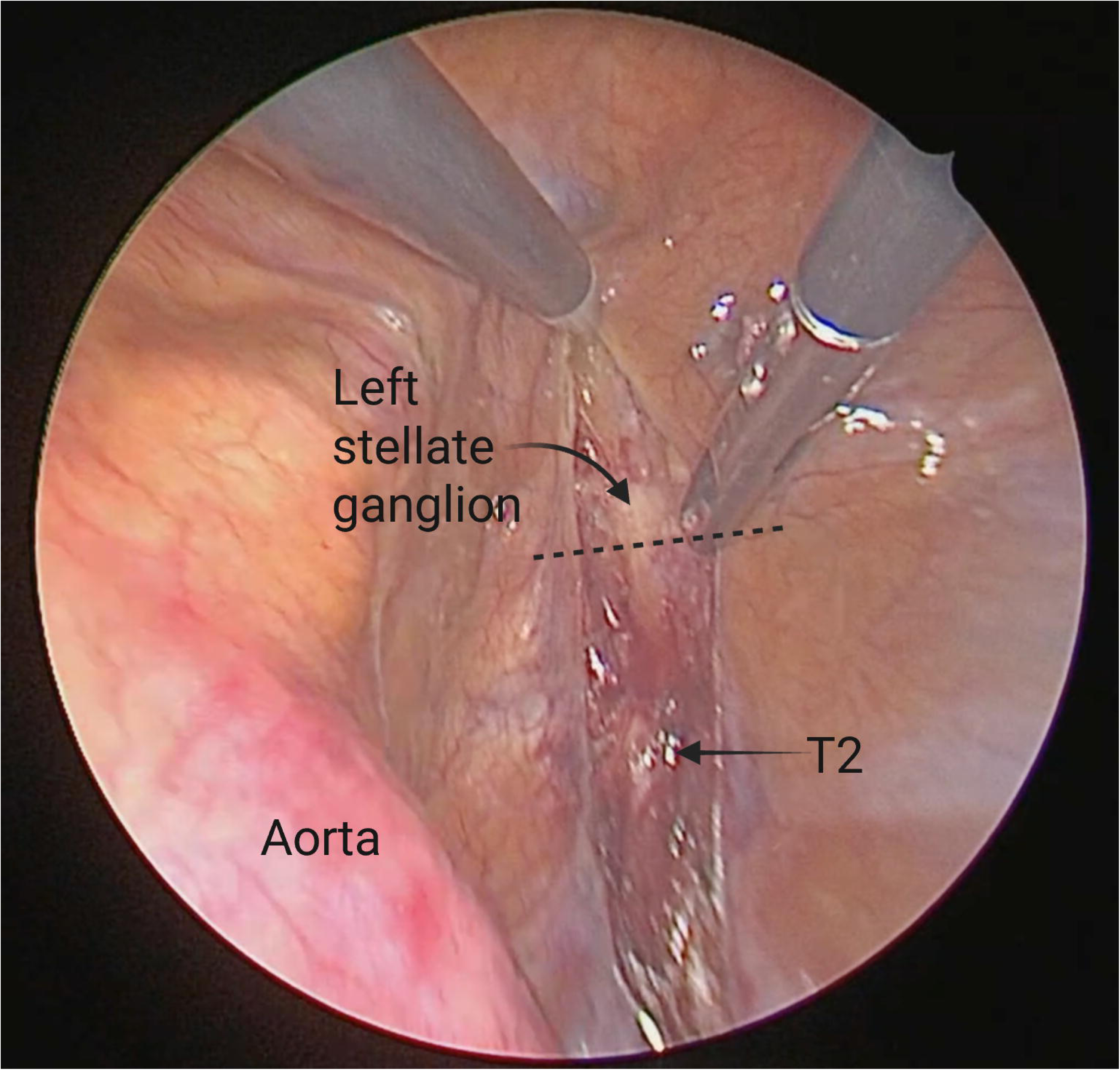
Thoracoscopic view exposing the left cardiac sympathetic chain. The left stellate ganglion and T2 are clearly visualized posterior and superior to the aortic arc. The sympathetic chain will be excised below the stellate ganglion until T4.

### Alternative Epicardial Access

A subxiphoid approach was performed via a 2–3 cm incision over the xiphoid-sternal joint, followed by dissection to the pericardium and insertion of a steerable sheath using the Seldinger technique or direct ablation using a cryoablation probe (Figure 1, Panel B). If needed, the xiphoid was removed to improve exposure.

In patients with adhesions and subxiphoid substrates, an inverted-T mini-sternotomy enabled adhesiolysis and targeted ablation. Alternatively, a limited anterolateral thoracotomy was used for anterior or LV summit substrates. A soft-tissue retractor (Alexis, Applied Medical, CA, USA) enhanced visualization and avoided rib spreading.

### V-A ECMO Support

In case of high PAINESD scores,^14^ ongoing VT, or preferred mapping during ongoing VT, V-A ECMO support was achieved by a small groin incision to expose the common femoral artery and vein. Purse-string sutures were placed, and cannulas were inserted using the Seldinger technique under echocardiographic guidance. The left groin was preferred to preserve the right side for endovascular EP access. Preoperative CT angiography was routinely performed to assess vascular anatomy. For patients anticipated to require rapid circulatory support, introducer sheaths were preemptively placed in the femoral vessels to allow expedited percutaneous cannulation if emergent ECMO initiation became necessary.

### Epicardial Mapping and Ablation

High-density epicardial mapping was performed using the CARTO system (Biosense Webster, Diamond Bar, CA, USA) with multipolar catheters (Pentaray, Octaray, or Optrell, Figure 3) or ablation catheters (ThermoCool, SmartTouch, or QDot). Bipolar electrograms were filtered at 10–400 Hz; scar was defined as peak-to-peak bipolar voltage <0.5 mV, and normal myocardium as >1.5 mV. Fractionated, late, and functionally delayed potentials^15^ were systematically annotated. If available, three-dimensional CCTA or CMR-derived scar models, including coronary or bypass trajectories, were merged into the EAM (Figure 3).

**Figure 3.**
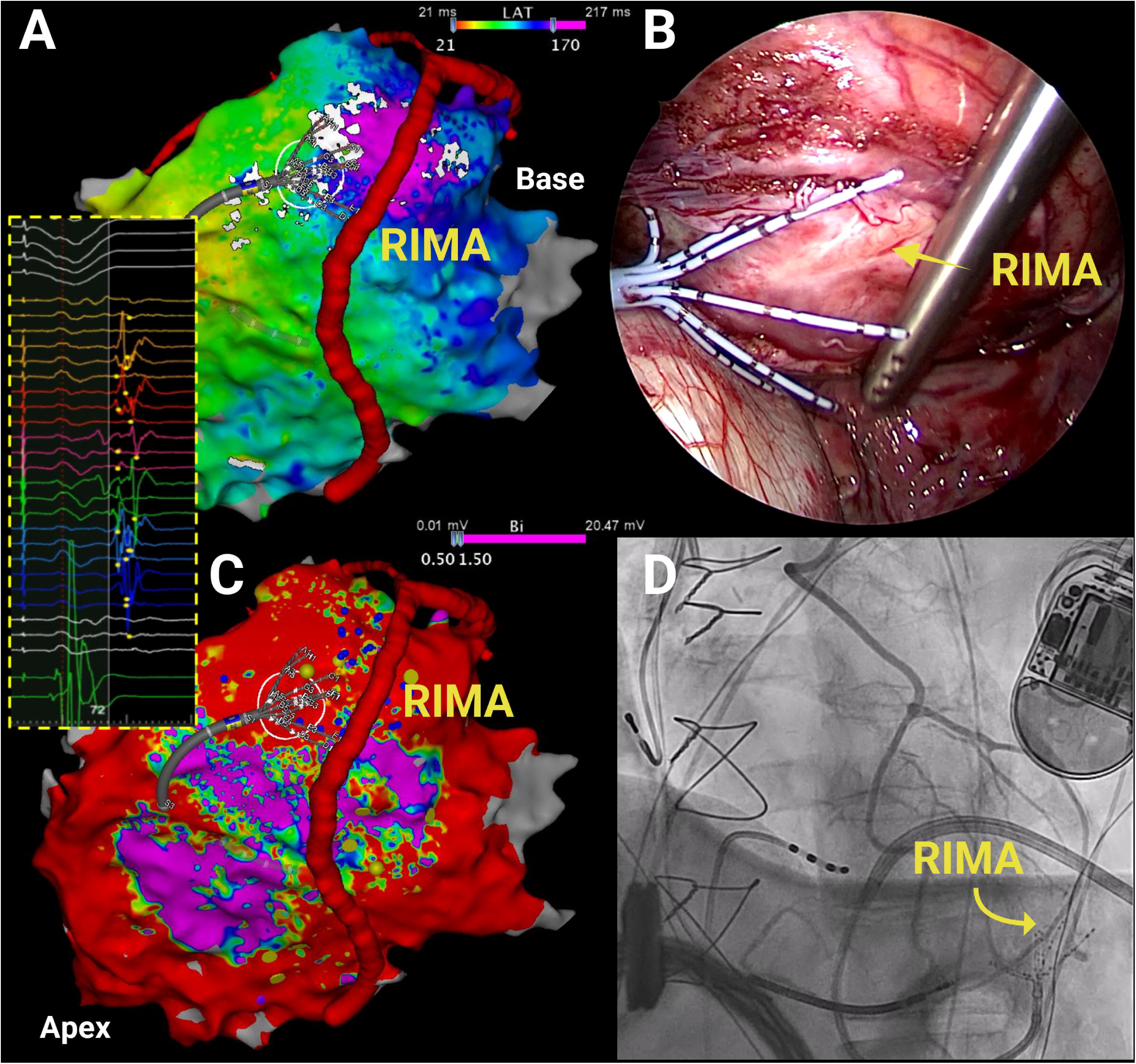
Epicardial co-localization of a multi-electrode mapping catheter to T-graft bypass. In Panel A, the epicardial position of the Octaray catheter in an area of late potentials (see inset) near the right-internal mammary artery (RIMA) bypass graft is visualized by a co-registered cardiac CT angiography reconstruction. Panel B, thoracoscopic confirmation of the position of Octaray covering the RIMA. Panel C, epicardial areas of reduced peak-to-peak voltage corresponding to delayed activation. Panel D, angiogram confirming the co-localization of the Octaray and the RIMA graft. Figure modified from thesis Darma, Maastricht University, 2025. RIMA = right-internal mammary artery.

Programmed electrical stimulation (PES; S2–S4 until refractoriness or 200 ms) was used to induce VT. Hemodynamically tolerated VTs were mapped using activation and entrainment; otherwise, substrate mapping was performed in sinus rhythm or during right ventricular pacing. Radiofrequency ablation (30–50 W, temperature 42 degrees Celsius) was targeted at areas with (evoked) delayed potentials, middiastolic activity, sites of concealed entrainment during VT, areas with >95% pace-map concordance (stim-to-QRS >30 ms), MRI-derived heterogeneous tissue channels,^12^ or CCTA-derived wall thickness channels.^11^ Coronary angiography and high-output pacing were performed before ablation to avoid coronary or phrenic injury.

Cryoablation was used when the arrhythmogenic substrate was within 10 mm of coronary vessels or bypass grafts, or when large contiguous areas required ablation. PES was repeated at the operator’s discretion; any remaining inducible VTs were subsequently targeted. Pericardial or pleural drains were placed as indicated and port sites were closed.

### Endpoints and Follow-Up

Acute procedural success was defined as non-inducibility of any VT. When PES was not performed or baseline non-inducibility was present, complete elimination of (evoked) delayed potentials was considered a surrogate endpoint for success. Partial success indicated non-inducibility of the clinical VT, with continued inducibility of non-clinical VTs; failure was defined as persistence of the clinical VT.

The primary endpoint was VT burden and ICD shocks during one-year follow-up, assessed via ICD interrogation and electronic records. Secondary endpoints included all-cause mortality, cardiovascular death, repeat VT ablation or stereotactic arrhythmia radioablation (STAR), ventricular assist device implantation, or heart transplantation. Major complications were defined as adverse events requiring intervention or prolonged hospitalization. Length of post-procedural hospital stay was also recorded.

### Statistical Analysis

Normality was assessed using the Shapiro–Wilk test. Continuous variables are presented as mean±standard deviation (SD) if normally distributed or median (interquartile range [IQR]) for non-normally distributed values, and categorical variables as counts (percentages). Changes in VT burden and ICD shocks were analyzed using the Wilcoxon signed-rank test. Event-free survival for all-cause mortality was evaluated with the Kaplan–Meier method. Statistical significance was set at P<0.05. Analyses were performed using GraphPad Prism (version 10.0.1, GraphPad Software Inc.) and Julius.ai.

## Results

### Patient Demographics

Between 2014 and 2025, twenty-two consecutive patients underwent minimally invasive hybrid VT ablation at our center. Baseline characteristics are summarized in Table 1. The cohort included equal numbers of patients with ischemic and nonischemic cardiomyopathies. Most patients presented with VT storm, experiencing a median burden of 16.5 [IQR 9.5–37.0] VT and 2 [IQR 0–5] ICD shocks in the year preceding the procedure. The median PAINESD score was 13 [IQR 4-17], reflecting a high risk of peri-procedural hemodynamic decompensation.

**Table 1:** Patient Demographics.

|  | Patients n = 22 |
| --- | --- |
| Age, y, median [IQR] | 70.1 [57.7-73.6] |
| Sex, male, n (%) | 19 (86) |
| LVEF, %, mean (SD) | 37.0 (14.3) |
| NYHA I-II, n (%) | 17 (77) |
| NYHA III-IV, n (%) | 5 (23) |
| ICMP, n (%) | 11 (50) |
| NICMP, n (%) | 11 (50) |
| - Genetic | 3 (14) |
| - ARVC | 3 (14) |
| - Myocarditis | 2 (9) |
| - Idiopathic | 3 (14) |
| VT storm, n (%) | 15 (68) |
| Comorbidities, n (%) |  |
| - Coronary artery disease | 12 (55) |
| - Diabetes mellitus | 4 (18) |
| - Hypertension | 16 (73) |
| - Stroke | 4 (18) |
| - Atrial fibrillation | 10 (46) |
| PAINESD score, median [IQR] | 13 [4-17] |
| Cardiac implantable electronic device, n (%) |  |
| - None | 1 (5) |
| - ICD | 18 (82) |
| - CRTD | 3 (14) |
| No. of prior endocardial VT ablations, mean (SD) | 1.5 (0.9) |
| Indications for hybrid ablation, n (%) |  |
| - Prior CABG | 5 (23) |
| - Extensive scar formation | 3 (14) |
| - Failed percutaneous epicardial access | 2 (9) |
| - Concomitant sympathetic denervation | 2 (9) |
| - Concomitant hybrid AF ablation | 1 (5) |
| - Prior hybrid AF ablation | 1 (5) |
| - Prior sternotomy for traumatic left-atrial |  |
| appendix rupture | 1 (5) |
| - Prior tamponade | 1 (5) |
| - Pericarditis | 1 (5) |
| - Preferred ablation under direct vision | 5 (23) |
| Medication, n (%) |  |
| - Beta-blocker | 17 (77) |
| - Amiodarone | 15 (68) |
| - Mexiletine | 0 (0) |
| - Sotalol | 3 (14) |
| ICD interrogation, median [IQR] |  |
| - VT burden | 16.5 [9.5-37.0] |
| - No. of ICD shocks | 2 [0-5] |

AF = atrial fibrillation; ARVC = arrhythmogenic right ventricular cardiomyopathy; CABG = coronary bypass grafting; CRT-D = cardiac resynchronization therapy defibrillator; ICD = implantable cardioverter-defibrillator; ICMP = ischemic cardiomyopathy; IQR = interquartile range; LVEF = left ventricular ejection fraction; NICMP = nonischemic cardiomyopathy; NYHA = New York Heart Association; SD = standard deviation; VT = ventricular tachycardia.

**Table 2:** Procedural Characteristics.

|  | Patients<br>n = 22 |
| --- | --- |
| Operation room, n (%) |  |
| - Hybrid EP operating room | 22 (100) |
| Surgical access, n (%) |  |
| - Left-sided VATS | 13 (59) |
| - Right-sided VATS | 1 (5) |
| - Subxiphoid surgical window | 3 (14) |
| - Combined left-sided VATS and subxiphoid surgical window | 2 (9) |
| - Anterolateral minithoracotomy | 3 (14) |
| No. of induced VTs, mean (SD) | 1.4 (1.0) |
| Pericardial adhesions, n (%) | 10 (45) |
| Epicardial ablation, n (%) | 22 (100) |
| Endocardial ablation, n (%) | 9 (41) |
| V-A ECMO support, n (%) | 5 (23) |
| Left-cardiac sympathetic denervation, n (%) | 2 (9) |
| Hybrid atrial fibrillation ablation, n (%) | 1 (5) |
| Procedure time, minutes, mean (SD) | 312 (98) |
| Acute outcome, n (%) |  |
| - Complete success | 16 (73) |
| - Partial | 1 (5) |
| - Failure | 0 (0) |
| - Not tested | 5 (23) |

EP = electrophysiology; SD = standard deviation; VATS = video-assisted thoracoscopic surgery; V-A ECMO = veno-arterial extracorporeal membrane oxygenation; VT = ventricular tachycardia

**Table 3:** Efficacy and Outcome of Minimally Invasive Hybrid VT Ablation.

|  |  |
| --- | --- |
| Days till discharge, days, median [IQR] | 7 [4-20] |
| VT recurrence, n (%) | 10 (45) |
| ICD interrogation, median [IQR] |  |
| - VT burden | 0 [0-5.8] |
| - No. of ICD shocks | 0 [0-0] |
| Repeat endocardial ablation, n (%) | 2 (9) |
| STAR, n (%) | 2 (9) |
| Acute complications, n (%) |  |
| - Hemothorax | 1 (5) |
| - Pericarditis | 2 (9) |
| Cardiovascular death, n (%) | 1 (5) |
| All-cause death, n (%) | 4 (18) |
| Heart transplantation, n (%) | 0 (0) |
| LVAD, n (%) | 0 (0) |

ICD = implantable cardioverter-defibrillator; IQR = interquartile range; LVAD = left-ventricular assist device; STAR = stereotactic arrhythmia radiotherapy; VT = ventricular tachycardia

Primary indications for minimally invasive hybrid ablation included prior coronary artery bypass grafting (CABG; n=5, 23%), extensive epicardial scarring (n=3, 14%), concomitant sympathetic denervation (n=2, 9%) or hybrid atrial fibrillation (n=1, 5%), failed percutaneous epicardial access (n=2, 9%), pericarditis (n=1, 5%), tamponade (n=1, 5%), prior sternotomy due to traumatic atrial rupture (n=1, 5%), and because of ablation under direct visualization (n=5, 23%). The mean left ventricular ejection fraction was 37±14%. Antiarrhythmic therapy before ablation included amiodarone or sotalol in 82% and beta-blockers in 77%. 20 patients had previously undergone an endocardial ablation (mean 1.5 ± 0.9 procedures).

### Minimally Invasive Epicardial Access

All hybrid procedures were performed in a dedicated hybrid EP operating room equipped for electrophysiological and surgical interventions. Most patients underwent a minimally invasive surgical approach using left-sided VATS (n=13, 59%). A subxiphoid surgical window was the primary access route in three patients (14%), and combined left-sided VATS with subxiphoid access in two cases (9%) for optimized epicardial exposure. A right-sided VATS was selected for one patient with arrhythmogenic right ventricular cardiomyopathy. V-A ECMO support was provided in five patients (23%) with a mean PAINESD of 17.

### Electrophysiologic Mapping and Ablation

Mapping was conducted with an ablation catheter before 2019 and with multielectrode catheters thereafter (Pentaray n=9, Octaray n=2, Optrell n=2). In six procedures, ablation was guided by integration of preprocedural CCTA or CMR-derived three-dimensional cardiac models (Figure 3). Programmed electrical stimulation induced a mean of 1.4±1.0 distinct VTs per patient.

The mean total procedure duration was 312±98 minutes. In most cases, initial mapping was performed with the pericardium intact, followed by opening and direct ablation under visualization (*ablation à vue*, Figure 1 Panels C and D). Epicardial ablation was performed in all patients, with adjunctive endocardial ablation in 50% (transseptal/transvenous in 5, retrograde aortic in 6). Radiofrequency energy was mostly used, with no occurrence of steam pops. In five patients (22%), ablation sites were adjacent to major coronary vessels or involved extensive epicardial substrate, prompting adjunctive cryoablation using a linear cryocatheter (Figure 1, Panel D). Pericardial adhesions were present in 45% of cases.

### Outcomes

#### Acute Procedural Outcomes

Complete procedural success was achieved in 16 patients (73%), partial success in one; in five patients, post-procedural programmed electrical stimulation was not performed because of frail patient status. No acute complications occurred. The median number of days until discharge was 7 days [IQR 4–20].

#### Follow-Up Outcomes

Within the follow-up period, the median VT burden decreased significantly to 0 [IQR 0–5.5]; P<0.001, representing an 81% reduction (Figure 4, Panel A). Similarly, the median number of ICD shocks decreased to 0 [0–0]; P<0.001, corresponding to a 90% reduction (Figure 4, Panel B). VT recurrences were observed in 45% of patients (Supplemental Figure 1) after one year. The Kaplan–Meier–estimated one-year all-cause survival rate was 82% (Figure 5).

**Figure 4.**
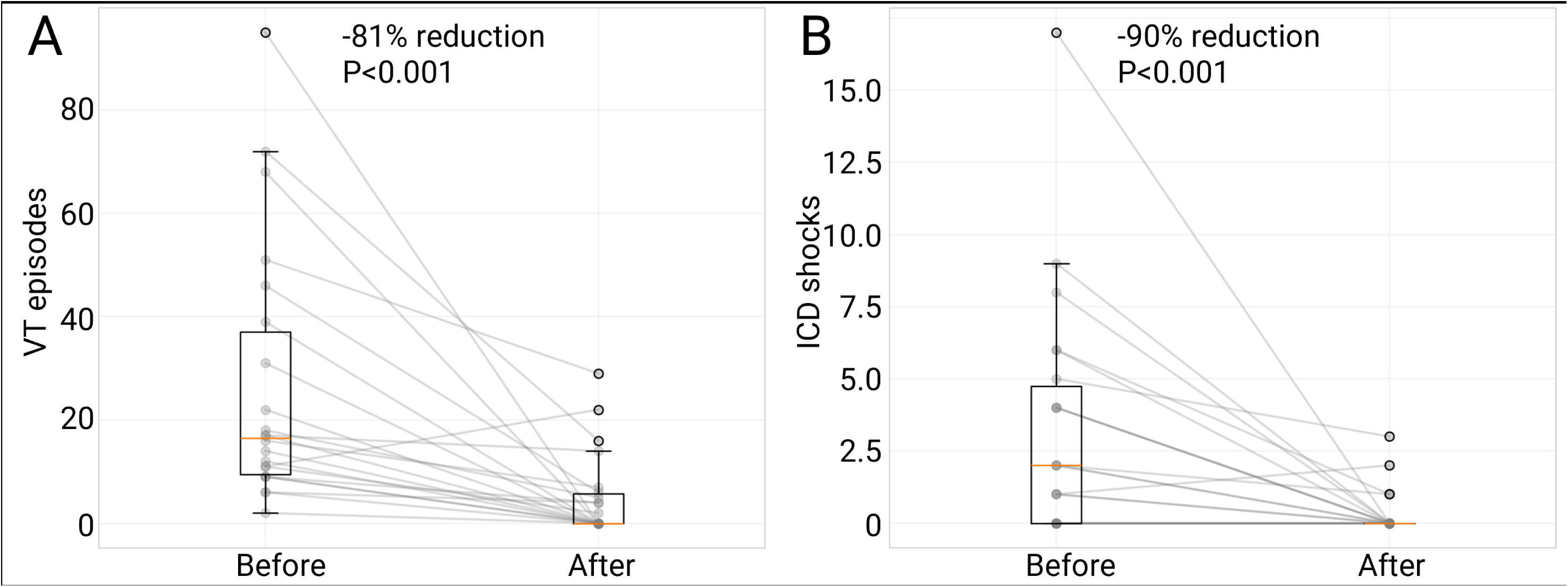
**Reduction of ventricular tachycardia episodes and internal cardioverter-defibrillator shocks**. Significant 81% and 90% reductions for median VT burden (16.5 [9.5-37.0] to 0 [0-5.8]; Panel A) and number of ICD shocks (2 [0-5] to 0 [0-0]; Panel B) were noted one year after minimally invasive hybrid VT ablation. ICD = internal cardioverter-defibrillator; VT = ventricular tachycardia.

**Figure 5.**
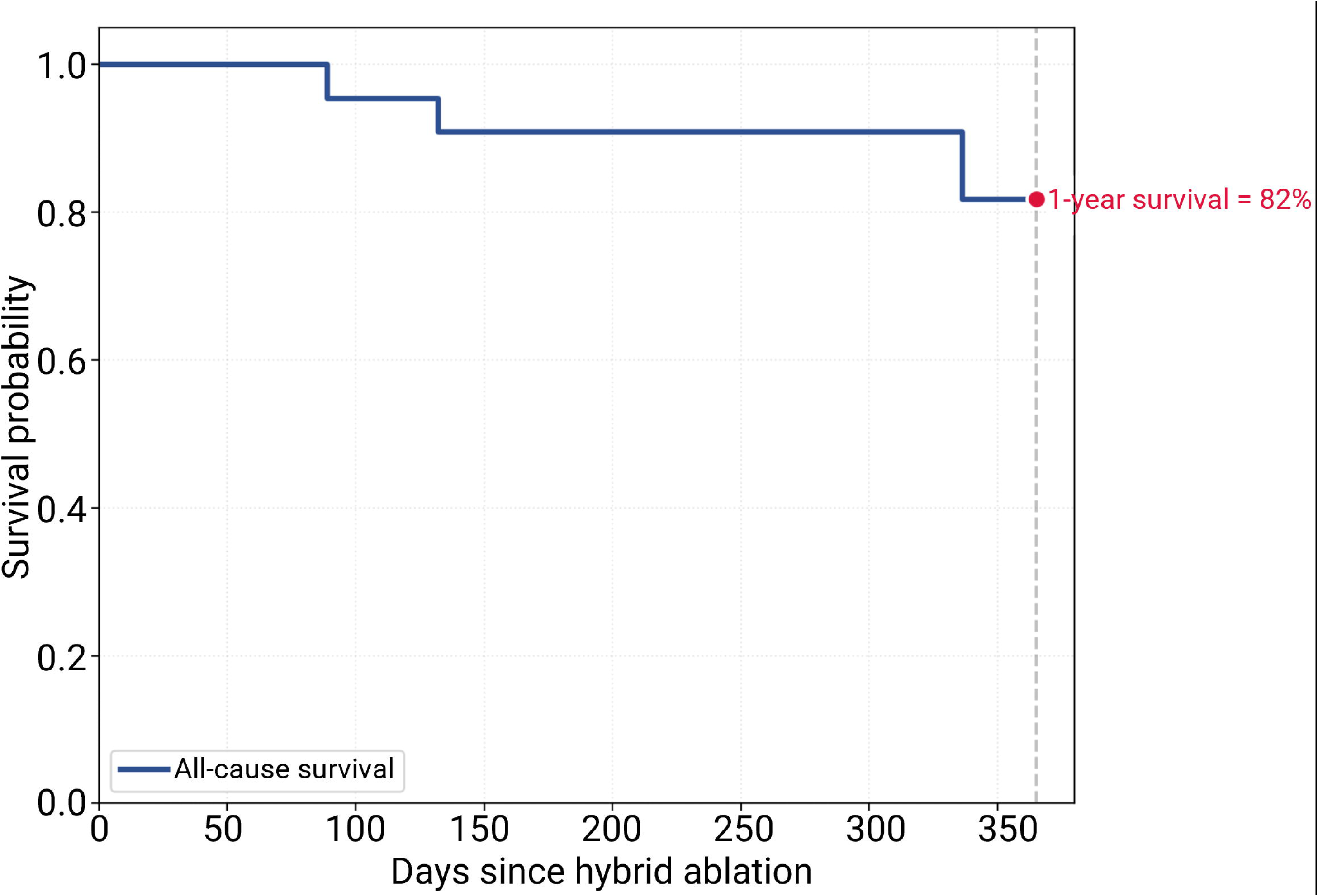
Kaplan-Meier curve of all-cause survival probability. A one-year survival of 82% was noted. No procedure-related deaths occurred. Four patients died: one from metastatic prostate cancer (day 89), one from an unknown cause (day 132), one from progressive heart failure (day 336), and one from respiratory failure during Pseudomonas bacteremia (day 336).

Four patients underwent repeat ablation procedures—radiofrequency catheter ablation (n=2), STAR (n=2). Subgroup analyses were not performed due to the limited sample size.

One major complication occurred—a hemothorax requiring blood transfusion and surgical exploration, without identification of an active bleeding source. Two patients developed postoperative pericarditis, both successfully treated with acetylsalicylic acid.

Four patients died within one year following hybrid ablation: one from metastatic prostate cancer (day 89), one from an unknown cause (day 132), one from progressive heart failure (day 336), and one from respiratory failure during Pseudomonas bacteremia (day 336).

## Discussion

This study represents the largest single-center experience of minimally invasive hybrid ablation for VT. Most patients underwent a left-sided VATS approach, enabling epicardial mapping, adhesiolysis, and substrate-specific ablation using radiofrequency or cryothermal energy. This approach uniquely allows for phrenic nerve mobilization, direct visualization of target sites (*ablation à vue*), and concomitant cardiac sympathetic denervation or hybrid atrial fibrillation ablation when indicated. Such complex procedures are optimally performed in a hybrid electrophysiology-surgical suite equipped to facilitate immediate escalation to cardiopulmonary support or open surgical intervention. Employing this patient-tailored, multidisciplinary strategy, we achieved procedural feasibility with significant reductions in VT burden and ICD shocks in a cohort characterized by advanced structural heart disease and substantial comorbidity burden (Central Illustration).

Depending on the underlying cardiomyopathy, the VT circuit often involves a complex three-dimensional substrate extending transmurally through the ventricular wall. Epicardial involvement is observed in up to 80% of ischemic cases and 77% of nonischemic cardiomyopathies,^4^ underscoring the need for epicardial access. Since the percutaneous subxiphoid approach was first introduced by Sosa et al in 1996,^16^ it has become the standard access for epicardial VT ablation.^7^ However, a successful route through the space of Larrey requires considerable expertise and carries a higher risk of complications (1-15%), even in high-volume centers.^17,18^ Furthermore, pericardial adhesions may be encountered in patients with prior cardiac surgery, pericarditis, or epicardial interventions. Percutaneous epicardial ablation after cardiac surgery could be performed safely in a small series;^19^ however, dense anterior adhesions frequently precluded effective mapping and ablation near bypass grafts. In a larger cohort of 18 post-surgical patients, Killu et al. reported a 67% success rate for percutaneous access, long procedure times, and several complications—including hemoperitoneum and ventricular lacerations—highlighting the limitations of this approach in patients with scarred pericardia.^20^

To improve safety and visualization in patients with prior cardiac surgery or failed percutaneous access, limited surgical windows were introduced. Soejima et al. demonstrated the feasibility of a subxiphoid surgical window in six patients, although mapping was often restricted to the inferior wall.^8^ Subsequently, Maury et al. and Michowitz et al. proposed tailored surgical access, either subxiphoid^21^ or limited anterior thoracotomy^21,22^, to improve epicardial exposure and facilitate direct ablation under vision. A subxiphoid surgical window (n=11; 3 partial sternotomy, 2 excision of xiphoid process for better exposure) preferentially provided access to the inferior and inferolateral epicardial walls, whereas a limited anterior thoracotomy (n=3) exposed the anterior, lateral, and apical walls. These studies demonstrated VT elimination in 50–70% of patients but also highlighted prolonged procedure times and significant complications, such as ventricular lacerations, underscoring the need for hybrid operating environments. In a multicenter cohort, Li et al. confirmed comparable efficacy between surgical and percutaneous epicardial access but again noted major complications in approximately 20% of patients.^9^

Despite its minimally invasive nature, current experience with thoracoscopic VT ablation in patients with structural heart disease remains limited. Aksu et al. first described successful VATS-guided ablation in refractory infero-lateral VT after multiple failed endocardial and epicardial procedures.^23^ In 2019, we reported a pilot series of five patients demonstrating the feasibility of minimally invasive hybrid VT ablation.^10^ The present study extends prior experience to 22 patients with ischemic and nonischemic cardiomyopathy, demonstrating that a structured, multidisciplinary selection and planning process in a tertiary center with expertise in both complex VT ablation and thoracoscopic procedures is safe and yields significant reductions in VT burden, even among high-risk patients.

Most patients underwent a left-sided VATS approach (59%), while a subxiphoid window was used in 14%. In selected cases, a double-sided (combined thoracoscopic and subxiphoid) access was required to achieve complete epicardial substrate coverage. This strategy is consistent with the recent report by Romero et al.^24^ Pericardial adhesions were present in 45% and bluntly dissected under direct vision. All patients underwent epicardial ablation, with additional cryoablation in five cases because of proximity to coronary arteries, extensive scarring, or epicardial fat. Concomitant cardiac sympathetic denervation was performed in two patients, a unique capability of the left-sided VATS, particularly relevant for patients with nonischemic cardiomyopathy or complex reentry circuits where sympathetic modulation may augment efficacy.^25^

Our results demonstrate an acute procedural success rate of 73% with a significant reduction in VT burden (−81%) and ICD shocks (−90%), and a 55% one-year VT-free survival rate. Only one hemothorax requiring reoperation occurred; no other major complications were observed. The one-year all-cause mortality of 18% reflects the high-risk nature of this cohort rather than procedure-related events. Ongoing technological advances, including the visualization of surgical ablation tools within mapping systems, are expected to further enhance procedural precision and safety.^26^

## Limitations

This retrospective study consists of highly selected cases referred to a tertiary hospital for catheter ablation of complex VT substrates and thoracoscopic surgery performed by operators with vast levels of expertise. Therefore, results on procedural efficacy and safety may not be generalizable to other electrophysiology laboratories.

## Conclusions

This study demonstrates that minimally invasive hybrid VT ablation using VATS access is a feasible option for patients with advanced structural heart disease, prior cardiac surgery, or failed percutaneous access. A multidisciplinary, patient-tailored approach enables epicardial mapping, controlled adhesiolysis, and direct visualization of complex substrates, with the unique possibility of concomitant left cardiac sympathetic denervation. The significant reduction in VT burden and ICD therapies with low procedural morbidity supports this hybrid strategy as a valuable addition to current VT ablation paradigms, warranting further prospective evaluation.

## Clinical Perspective

Minimally invasive hybrid VT ablation using VATS access offers a feasible treatment option for patients with advanced structural heart disease, prior cardiac surgery, or failed percutaneous epicardial access. This approach allows direct visualization of the epicardial substrate, controlled adhesiolysis, and the option for concomitant cardiac sympathetic denervation, resulting in substantial reductions in VT burden and ICD therapies.

## Translational Outlook

Future prospective and multicenter studies should compare thoracoscopic hybrid ablation with conventional percutaneous strategies to better define patient selection criteria, procedural standardization, and long-term clinical outcomes. Integration of real-time imaging and mapping technologies may further improve procedural precision and safety.

## Data Availability Statement

Data will be shared upon reasonable request by the corresponding author (RtB).

## Data Availability

All data produced in the present study are available upon reasonable request to the authors

## Acknowledgments

Illustrations were partly created with a licensed version of BioRender.com

## Tweet

Hybrid thoracoscopic VT ablation enables epicardial mapping, direct visualization, and tailored substrate ablation with low complication rates. A multidisciplinary strategy yields durable arrhythmia suppression in high-risk patients. #JACCClinEP #Electrophysiology #VTablation

## Abbreviations

ARVC: Arrhythmogenic right ventricular cardiomyopathy
CABG: Coronary artery bypass grafting
CCTA: Cardiac computed tomography angiography
CMR: Cardiac magnetic resonance imaging
CRT-D: Cardiac resynchronization therapy defibrillator
EAM: Electroanatomic map
VA-ECMO: Veno-arterial extracorporeal membrane oxygenation
EP: Electrophysiology
ICD: Implantable cardioverter-defibrillator
ICMP: Ischemic cardiomyopathy
LCSD: Left-cardiac sympathetic denervation
LVEF: Left ventricular ejection fraction
NICMP: Nonischemic cardiomyopathy
NYHA: New York Heart Association
PES: Programmed electrical stimulation
STAR: Stereotactic arrhythmia radioablation
VATS: Video-assisted thoracoscopic surgery
VT: Ventricular tachycardia

---

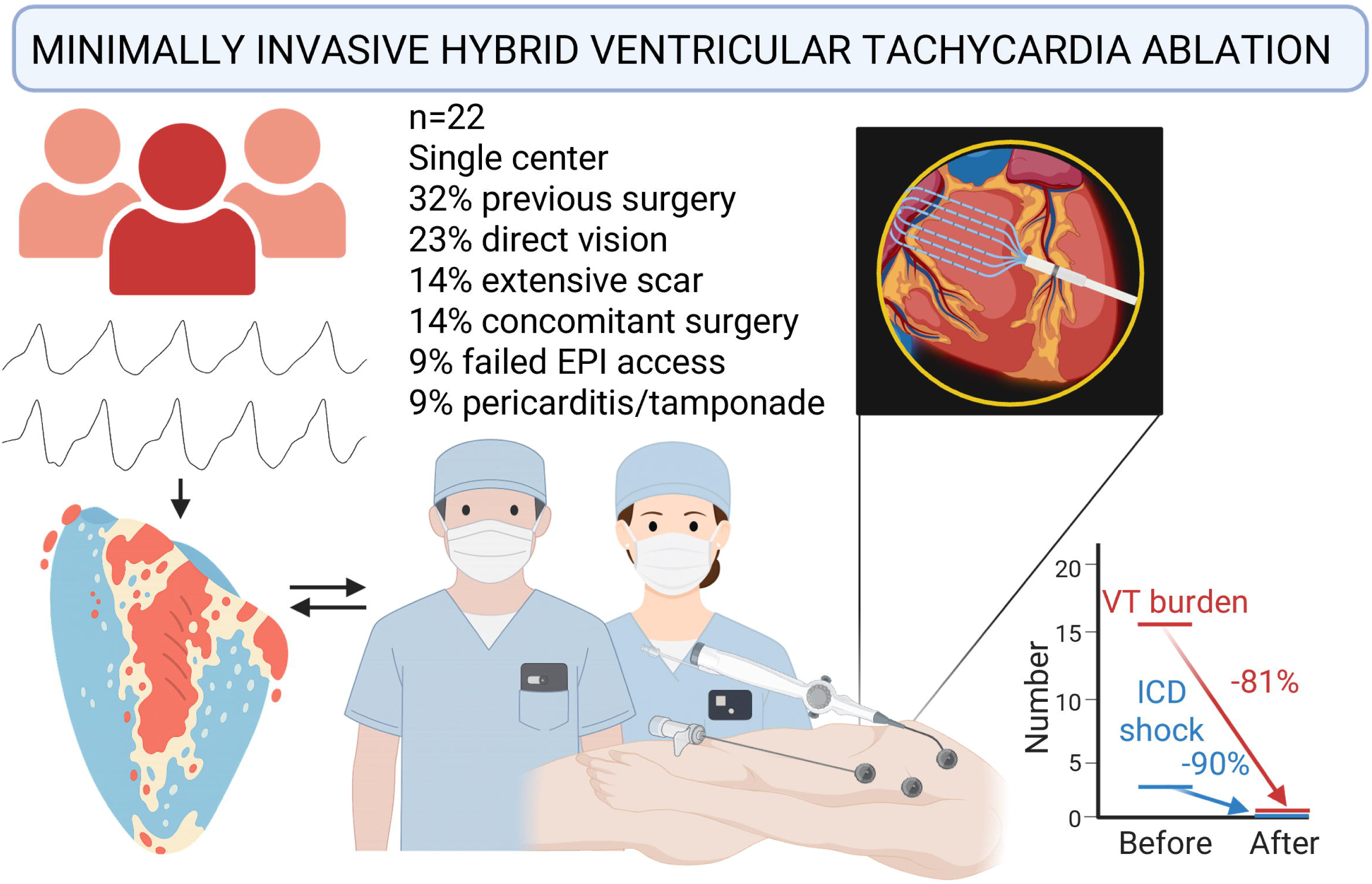
Central Illustration. Minimally invasive hybrid ventricular tachycardia ablation. In this study, following a comprehensive preoperative evaluation within the structured VT-TRACT care pathway, most eligible patients underwent a left-sided video-assisted thoracoscopic hybrid VT ablation. This approach allowed for phrenic nerve mobilization, direct visualization of target sites (ablation à vue), and concomitant cardiac sympathetic denervation. Employing this patient-tailored, multidisciplinary strategy, we achieved high procedural efficacy and safety in a cohort characterized by advanced structural heart disease and substantial comorbidity burden.

