## Supplementary figures and images for "Patient-Tailored Minimally Invasive Hybrid Ablation of Complex Ventricular Tachycardia Substrates"

### Supplemental Figure 1

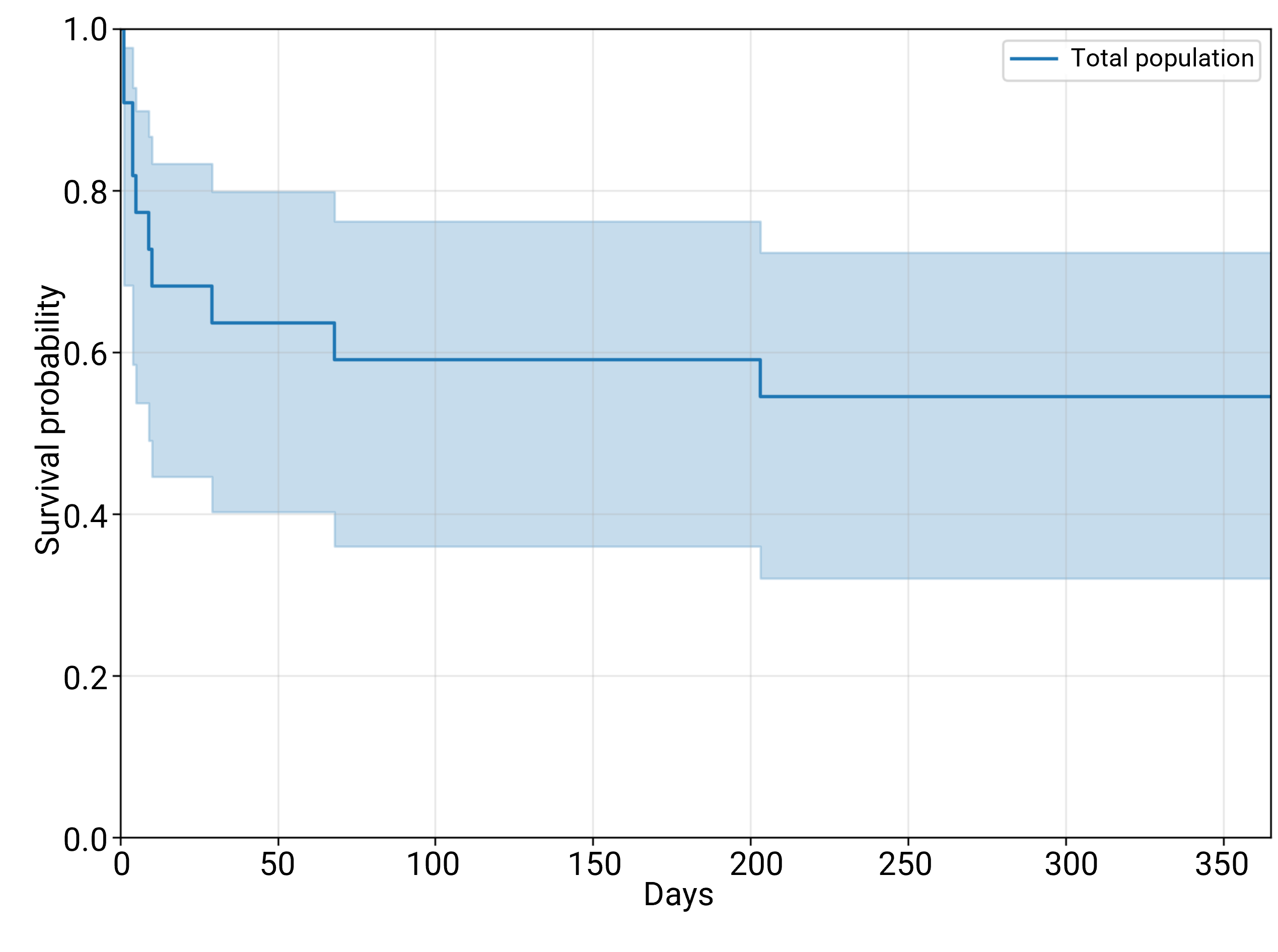
